# Diabetes testing, care cascade and the burden of undiagnosed diabetes in Kenya. Evidence from the Kenya STEPwise Survey

**DOI:** 10.64898/2026.09.29.26364371

**Authors:** Catherine Akoth, Chloe V. Thomas, James Odhiambo Oguta, Elvis Omondi Achach Wambiya, Peter J. Dodd

**Affiliations:** Sheffield Centre for Health and Related Research (SCHARR), Division of Population Health, School of Medicine and Population Health, University of Sheffield, United Kingdom; Centre for Health Economics and Decision Modelling, Health Economics and Decision Science Institute (HEDSI), Nairobi, Kenya; Chronic Disease Management Unit, African Population and Health Research Center (APHRC), Nairobi, Kenya

**Keywords:** Diabetes, screening, diagnosis, testing, care cascade, undiagnosed diabetes

## Abstract

**Background:** Diabetes is a growing public health challenge globally, with a disproportionate burden occurring in low- and middle-income countries (LMICs). In Kenya, limited evidence exists on type 2 diabetes mellitus (T2DM) screening coverage, undiagnosed diabetes, and progression through the diabetes care cascade. This study assessed the prevalence and determinants of T2DM testing, quantified undiagnosed T2DM, and examined the T2DM care cascade in Kenya.

**Methods:** We analysed data from the 2015 Kenya WHO STEPwise survey, a nationally representative cross-sectional survey of adults aged 18–69 years. The analysis included 4,167 participants after excluding individuals with missing fasting plasma glucose (FPG) measurements and those below the age of 18 and above 69 years. Diabetes testing was defined as self-reported prior blood glucose measurement by a health worker among individuals without known diabetes. Diabetes was defined as FPG ≥7 mmol/L, previous diagnosis, or current diabetes treatment. Weighted prevalence estimates with 95% confidence intervals (CI) were calculated. Univariable and multivariable logistic regression analyses were conducted to identify factors associated with diabetes screening while accounting for the complex survey design.

**Results:** Overall, 11.9% (95% CI: 9.7–14.5) of participants reported ever being tested for diabetes. The weighted prevalence of diabetes was 2.66% (95% CI: 2.06–3.42), while 53.4% (95% CI: 39.0–67.4) of diabetes cases were previously undiagnosed. Testing prevalence was highest among older adults, individuals with tertiary education, obese participants, and those with hypertension, but lowest among younger adults, individuals with no formal education, and residents of the North Eastern region. Among individuals with diabetes, 46.9% were aware of their diagnosis, 40.8% of those aware were on treatment, and only 26.4% of those on treatment had achieved glycaemic control. In multivariable analysis, older age (60–69 years: aOR=6.29, 95% CI: 3.66–10.79), tertiary education (aOR=2.65, 95% CI: 1.33–5.27), poorest wealth quintile (aOR=8.42, 95% CI: 4.11–17.24), obesity (aOR=2.13, 95% CI: 1.41–3.21), and hypertension (aOR=1.64, 95% CI: 1.21–2.23) were significantly associated with increased odds of diabetes testing.

**Conclusion:** Diabetes testing coverage in Kenya remains low, with more than half of diabetes cases remaining undiagnosed and substantial losses occurring across the diabetes care cascade. These findings highlight critical gaps in early diagnosis, linkage to care, treatment uptake, and glycaemic control. Strengthening integrated diabetes screening and care services, particularly for underserved and high-risk populations, is essential to improve early detection and continuity of care in Kenya.

## Introduction

Diabetes is a growing global threat to public health, accounting for 9.3% of all-cause mortality, with more than 500 million people currently living with the disease(1). Type 2 diabetes mellitus(T2DM) alone accounts for 90% of all the cases(2). The burden of T2DM is increasing exponentially in low- and middle-income countries (LMICs), which also have more than 80% of all diabetes cases(1). The global trend is predicted to continue rising, with approximately 800 million expected to be living with diabetes by 2050(1).

Kenya, like other LMICs, is experiencing an epidemiological transition characterised by the rising burden of Non-Communicable Diseases (NCDs) (3) while infectious diseases also remain a constant threat(4–6). The rising burden has been attributed to dietary shifts to processed foods, urbanization and adoption of a sedentary lifestyle(7–9). T2DM is a major cause of morbidity and mortality in Kenya, with a prevalence of 2.4%(10). It is not only a health problem but also creates an economic burden to the health system, households and individuals(11). A defining aspect of the diabetes threat is the burden of undiagnosed diabetes, which results in silent progression of the disease, given that diabetes is mainly asymptomatic during the early stages(12). Undiagnosed diabetes, therefore, increases the risk of developing microvascular and macrovascular complications by the time it is detected(12). Globally, 40% of adults with diabetes are unaware of their condition(13,14), while in Kenya, more than half of people with diabetes remain undiagnosed(10). These high rates of undiagnosed diabetes denote a gap in diagnosis as a critical barrier to accessing care for diabetes.

The impact of diabetes can be addressed through various means. These include preventive interventions to prevent, delay the onset, or reverse the disease, as well as early diagnosis and initiation of care to manage glucose levels and prevent complications(2). Such interventions include lifestyle modification(15–17), diet and physical activity(2), and pharmacological interventions(18,19). Screening for diabetes is a key strategy for early identification of individuals with diabetes(20–22), especially in LMICs like Kenya where routine health services for chronic diseases are limited (23–26). Screening is done through blood sugar testing or risk assessment and can be done at different settings including the community and health facilities. Screening for diabetes in Kenya is mainly opportunistic rendering a large proportion of the population unreached with the necessary screening services(24,27). The World Health Organization (WHO) recommends the integration of diabetes screening into primary health care settings to facilitate care in high burden countries(28).

While screening is a key tool for initiating diabetes care, it is inadequate on its own. The continuum of care for diabetes within the health system, beyond screening, is essential for effective management of diabetes(20). The care cascade concept has been applied widely for infectious diseases and has been recently adopted for diabetes to quantify diagnosis and linkage to care, treatment initiation, and glycaemic control among people with diabetes(29–32). Understanding the care cascade beyond diagnosis is key for policymakers to ensure adequate response at each cascade stage.

Previous studies in Kenya have focused on the prevalence of diabetes and prediabetes with no emphasis on screening and the care cascade (10,33). This study, therefore, sought to determine the prevalence and factors associated with self-reported diabetes screening through blood sugar testing, the distribution of undiagnosed diabetes, and to construct and quantify the diabetes care cascade in Kenya. The findings from this study are expected to provide evidence for policymakers to design effective and equitable diabetes screening strategies and care across the cascade.

## Methods

### Data Source and Study Design

We analysed data from the 2015 Kenya WHO STEPwise survey, which is the first nationally representative cross-sectional household survey on NCD risk factors among adults aged 18-69 years, conducted across the 47 counties of Kenya between April and June 2015, using a standardized questionnaire(34). The study employed a multistage stratified sampling approach using the fifth National Sample Surveys and Evaluation Programme (NASSEP V) master sampling frame, developed by the Kenya National Bureau of Statistics based on enumeration areas from the 2009 Kenya Population and Housing Census (34). The frame comprised 5,360 clusters, divided into four equal subsamples, out of which 6,000 households were selected nationwide, with one eligible adult randomly selected per household. Among these, 4,754 households consented to participate, yielding 4,500 completed interviews. Further details of the survey design have been described in detail elsewhere(34). The 2015 Kenya WHO STEPwise Survey of Non-Communicable Disease Risk Factors was conducted following ethical approval from the Kenya Medical Research Institute (KEMRI) Ethics Review Committee and obtaining informed consent of all survey participants. The analysis was conducted using de-identified survey data, with no access to identifiable participant information. Therefore, additional ethical approval was not sought for the present secondary analysis.

### Measures

#### Outcome variables

In this study, the outcome variable, which is diabetes testing, was operationalised as a self-report of blood glucose measurement among individuals without known diabetes. It was assessed using the question: *Has your blood sugar ever been measured by a health worker?* For which the responses were *yes* or *no*. Individuals who reported a previous diabetes diagnosis by a health worker were excluded.

The diabetes care cascade was assessed based on three variables:

1. Diabetes, defined as having fasting plasma glucose (FPG) results of ≥7 mmol/L or a previous diabetes diagnosis by a health worker or ever having been treated for diabetes, and was categorised as *‘diabetes’*, otherwise ‘*no diabetes’*.
2. Awareness, defined as a self-report of ever having been diagnosed with diabetes by a health worker. This was assessed using the question: *Have you ever been told by a doctor that you have diabetes?* The responses were *‘yes’* or ‘*no’*.
3. Undiagnosed diabetes, defined as having diabetes but never having been told by a healthcare provider that they have diabetes. This variable is an inverse of the ‘*Awareness*’ variable above. It was assessed using the question: *Have you ever been told by a doctor that you have diabetes?* The responses were recoded as ‘*undiagnosed’* if response is *no* and ‘*diagnosed’* if response is *yes*.
4. Treatment, defined as a self-report of ever having received treatment for diabetes among those aware. This was assessed using the question: *Are you currently taking medication for diabetes?*
5. Glycaemic control, defined as FPG < 7 mmol/L among those receiving treatment.

#### Independent variables

The explanatory variables were selected based on their relevance to diabetes screening and their availability in the datasets. The following are the included variables and their corresponding categories: Sex (*male, female*), Age groups (*18-29 yrs, 30-44 yrs, 45-59yrs, 60-69 yr*s), Marital status (*married, single*), Education level (*no formal, primary, secondary, tertiary*), Occupation (*unemployed/unpaid, self-employed, employed/salaried*), Residence (*rural, urban*), Region (R*ift Valley, Coast, Nyanza, Western, Eastern, North Eastern, Nairobi, Central*). Principal component analysis of household assets was used to compute the wealth index (35,36) which was categorised into five wealth quintiles (*poorest, poorer, middle, richer, richest*). Health status variables included were obesity(*obese, not obese*) and hypertension (*hypertension, no hypertension*).

### Statistical analysis

Sample characteristics were summarised using frequencies and percentages. Prevalence of screening and undiagnosed diabetes were summarised using percentages and their 95% confidence intervals. Univariable and multivariable logistic regression analyses were performed to assess factors associated with diabetes screening. All the covariates in the bivariate model were included as confounders in the multivariable model. The model’s goodness of fit was assessed using the Hosmer-Lemeshow test(37). All analysis was conducted using Stata software version 19.5(38). Clustered sampling design was adjusted for using the survey weights provided in the dataset and the *svy* command in Stata.

## Results

### Sample Characteristics

Out of the 4500 sampled individuals, a total of 4167 adults aged 18-69 were included in the analysis, after excluding 14 individuals outside the age range and 319 observations with missing values for FPG measurements. Most of the participants in the sample were female(60.6%), were aged 30-44 years(38.2%), were married(68.2%) and had attained primary education(46.5%). A major proportion of the sample consisted of self-employed people(39.6%), in the richest wealth quintile(22.4%), and from the Rift Valley region(30.4%). The sample was almost evenly distributed between urban(51.3%) and rural(48.7%) residences. Ten percent of the participants were obese, while 24.7% were hypertensive. (**Table 1**)

**Table 1:**
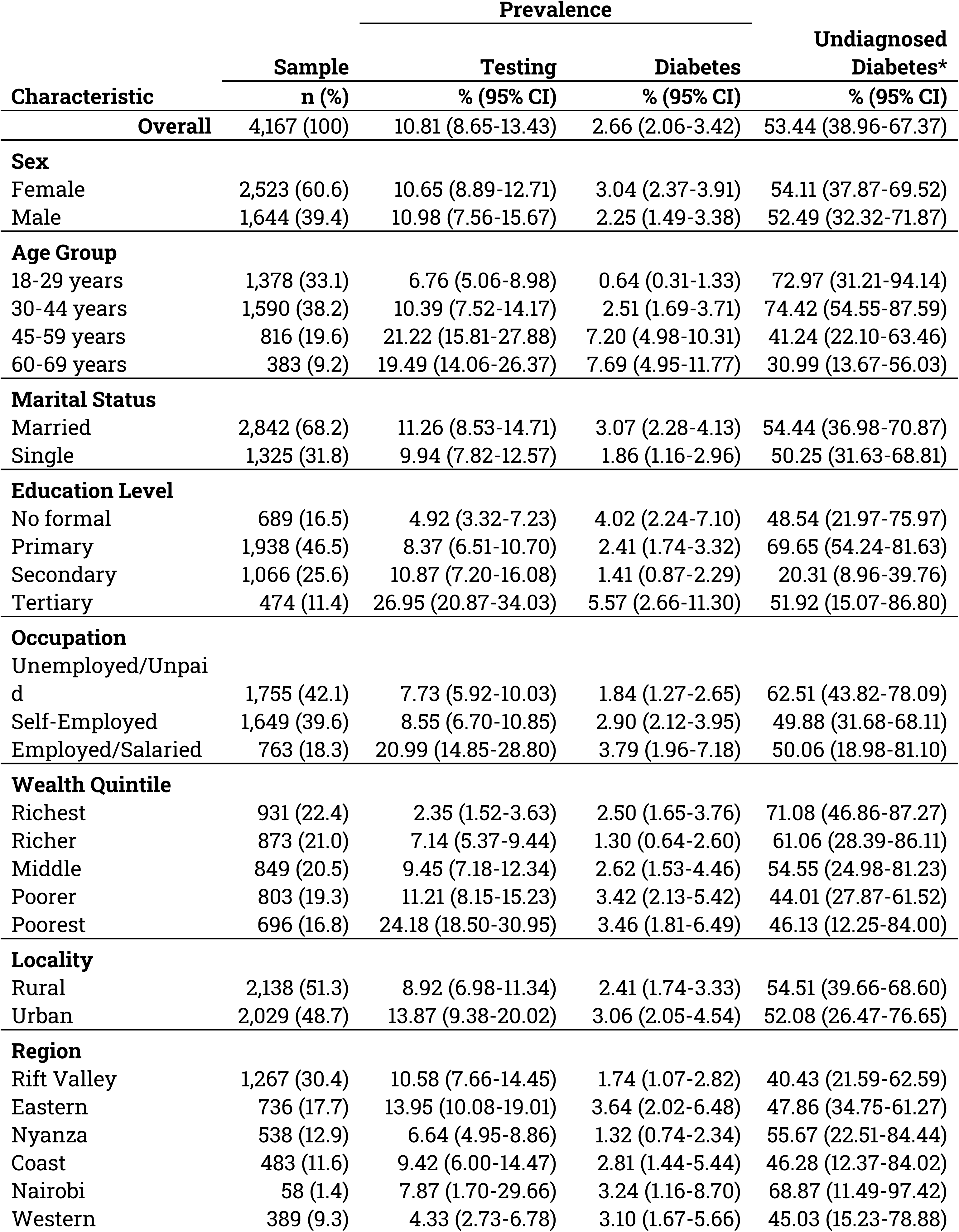
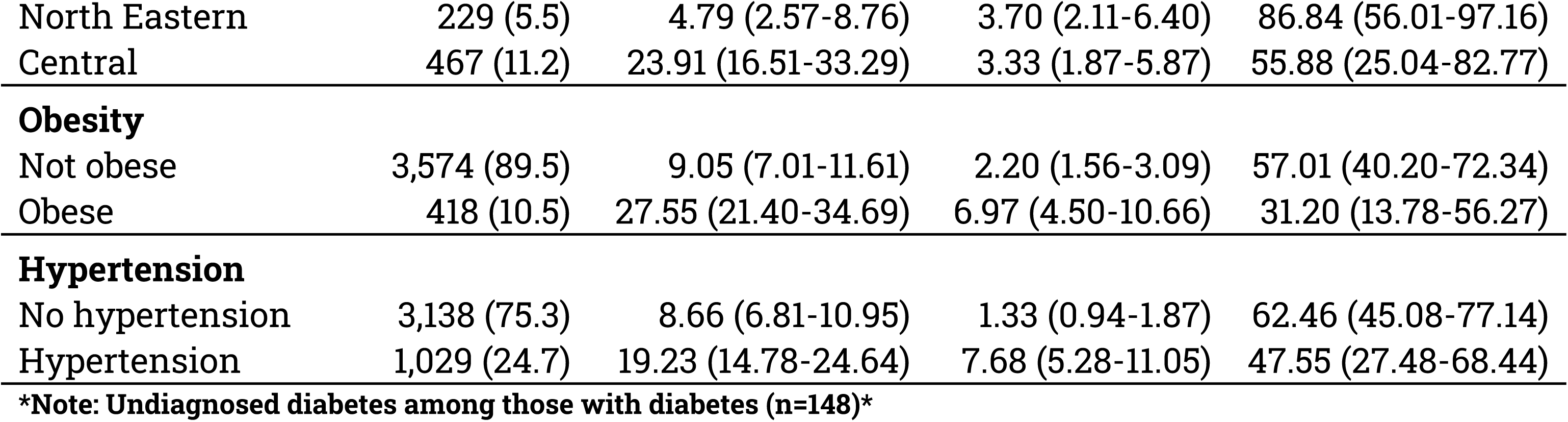
Sample characteristics, testing prevalence, diabetes prevalence and proportion of diabetes that is undiagnosed.

### Prevalence of testing for diabetes

Overall, only 10.81%(95% CI: 8.65-13.43) of the participants reported ever having been tested for diabetes. The prevalence was highest among older adults aged 45-59 years(21.2%) and 60-69 years(19.5%), those with tertiary education level(26.9%), urban residents(13.9%), those from the Central region(23.9%) and obese individuals(27.6%). Conversely, the prevalence was lowest among younger adults aged 18-29 years(6.8%), the uneducated(4.9%), the unemployed(7.7%), those in the richest wealth quintile(2.4%), and among those from the Northeastern region(4.8%). (**Table 1**)

### Prevalence of diabetes

The weighted prevalence of diabetes in Kenya was 2.66%(2.06-3.42) and was highest among older adults [45-59 years, 60-69 years (7.2%, 7.69%) respectively], married(3.07%), tertiary educated(5.57%), employed(3.79%), participants from Northeastern(3.7%), obese(6.97%) and hypertensive participants(7.68%).

### Proportion of undiagnosed diabetes

The proportion of undiagnosed diabetes was 53.4% (39.0-67.4). The proportion of undiagnosed diabetes was highest among younger adults aged 18-29 years(73%) and 30-44 years(74.4%), those with primary level education(69.7%), the unemployed(62.5%), those in the richest wealth quintile(71.1%), those from the Northeastern region(86.8%) and normotensives(62.5%). In contrast, lower undiagnosed rates were observed among older adults aged 60-69 years(31%), individuals with secondary education(20.3%) and obese individuals(31.2%). (**Table 1**)

### Testing coverage versus disease burden

Older adults aged 45-69 years had the highest testing coverage (24-25%), corresponding with the highest diabetes prevalence (7.2-7.7%) reported. Obese individuals reported more testing (31.0%) than non-obese (9.9%), matching their higher diabetes prevalence (6.97% vs. 2.20%). Similarly, hypertensives reported higher testing (22.5%) than non-hypertensives (9.1%), aligning with their higher prevalence (7.68% vs. 1.33%). The poorest wealth quintile reported the highest testing (25.6%), corresponding with the highest diabetes burden (3.46%), suggesting effective reach of public health services to these high-risk/ high burden groups. Adults with no formal education had high diabetes prevalence (4.02%) but among the lowest testing coverage (6.9%). The richest wealth quintile had only 3.1% testing coverage despite having high diabetes prevalence (2.50%). Regionally, Northeastern had the most severe misalignment with the highest diabetes prevalence (3.70%) yet the lowest testing in the country (5.3%). The under tested high burden groups reveal critical gaps where testing does not match the need. (**Table 1**)

### Diabetes care cascade

The weighted prevalence of diabetes was 2.66% (95%CI: 2.06-3.42). Among individuals with diabetes, less than half (46.96%, 95%CI: 32.63-6104) were aware of their diagnosis and out of those aware, only 40.8% (95%CI: 24.59-37.35) reported being on treatment for diabetes. Among those receiving treatment, only 26.39% (95%CI: 11.07-50.79) had achieved glycaemic control. (**Figure 1**)

**Figure 1.**
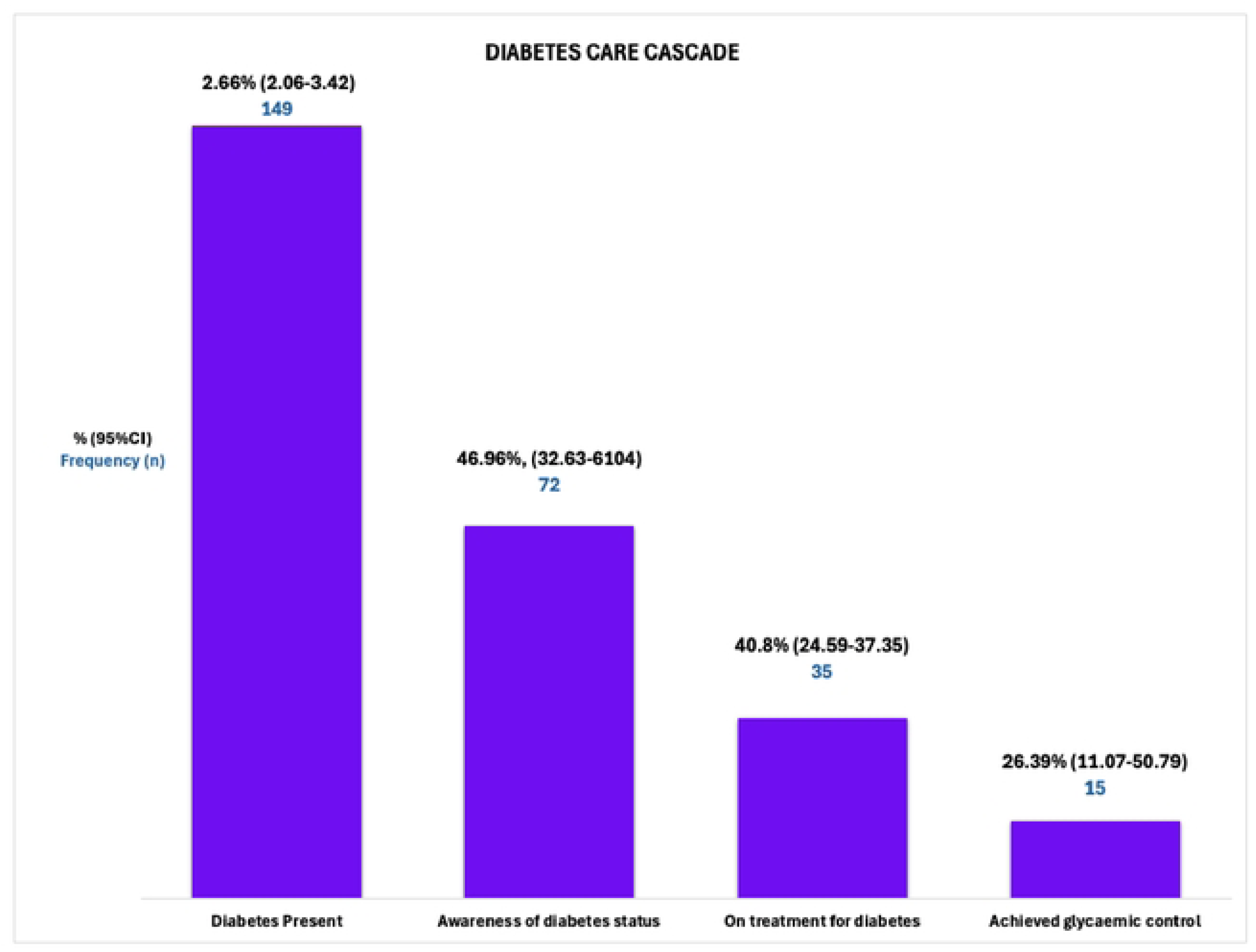
Diabetes care cascade

### Factors associated with diabetes screening

In the univariable logistic regression analysis, the odds of diabetes testing increased with increasing age: older adults (45-59 years: cOR=3.72, 95%CI: 2.56-5.40), (60-69 years: cOR=3.34, 95% CI:1.96-5.68) were more likely to be tested compared to younger adults. Tertiary education(cOR= 7.13, 95% CI: 4.20-12.10), poorest wealth quintile (cOR= 13.26, 95% CI: 7.52-23.38), the Central region (cOR= 2.66, 95% CI: 1.48-4.77), obesity (cOR= 3.82, 95% CI: 2.62-5.56), and having hypertension (cOR= 2.51, 95% CI: 1.86-3.39) were also associated with increased odds of being tested. The odds of testing increased with reducing wealth status. Nyanza (cOR= 0.60, 95% CI: 0.37-0.971), Western (cOR= 0.38, 95% CI: 0.21-0.69) and Northeastern (cOR= 0.43, 95% CI: 0.20-0.89) regions had lower odds of diabetes testing compared to the Rift Valley region. **(Table 2)**

**Table 2:** Factors associated with self-reported diabetes testing.

| Characteristics | Univariable logistic regression |  | Multivariable logistic regression |  |
| --- | --- | --- | --- | --- |
|  | cOR (95% CI) | p-value | aOR (95% CI) | p-value |
| **Sex** |  |  |  |  |
| Female | 1 |  | 1 |  |
| Male | 1.03 (0.69-1.56) | 0.871 | 0.78 (0.54-1.12) | 0.183 |
| **Age Group** |  |  |  |  |
| 18-29 years | 1 |  | 1 |  |
| 30-44 years | 1.60 (1.09-2.35) | **0.018** | 1.62 (1.04-2.54) | 0.033 |
| 45-59 years | 3.72 (2.56-5.40) | **<0.001** | 4.20 (2.63-6.71) | **<0.001** |
| 60-69 years | 3.34 (1.96-5.68) | **<0.001** | 5.57 (2.92-10.65) | **<0.001** |
| **Education** |  |  |  |  |
| No formal | 1 |  | 1 |  |
| Primary | 1.77 (1.10-2.84) | **0.019** | 1.61 (0.93-2.79) | 0.085 |
| Secondary | 2.36 (1.30-4.27) | **0.005** | 1.72 (0.91-3.26) | 0.094 |
| Tertiary | 7.13 (4.20-12.10) | **<0.001** | 3.58 (1.71-7.47) | **0.001** |
| **Wealth Quintile** |  |  |  |  |
| Richest | 1 |  | 1 |  |
| Richer | 3.20 (1.84-5.55) | **<0.001** | 2.95 (1.50-5.83) | **0.002** |
| Middle | 4.34 (2.63-7.14) | **<0.001** | 3.82 (2.02-7.20) | **<0.001** |
| Poorer | 5.25 (2.97-9.27) | **<0.001** | 4.07 (2.02-8.19) | **<0.001** |
| Poorest | 13.26 (7.52-23.38) | **<0.001** | 10.03 (4.77-21.09) | **<0.001** |
| **Residence** |  |  |  |  |
| Rural | 1 |  | 1 |  |
| Urban | 1.64 (0.98-2.75) | 0.059 | 1.10 (0.82-1.48) | 0.523 |
| **Marital Status** |  |  |  |  |
| Married | 1 |  | 1 |  |
| Single | 0.87 (0.62-1.23) | 0.43 | 1.26 (0.90-1.74) | 0.173 |
| **Region** |  |  |  |  |
| Rift Valley | 1 |  | 1 |  |
| Eastern | 1.37 (0.82-2.29) | 0.228 | 1.15 (0.70-1.88) | 0.578 |
| Nyanza | 0.60 (0.37-0.97) | **0.036** | 0.74 (0.45-1.21) | 0.235 |
| Coast | 0.88 (0.48-1.61) | 0.674 | 0.80 (0.48-1.31) | 0.373 |
| Nairobi | 0.72 (0.14-3.71) | 0.694 | 0.46 (0.16-1.30) | 0.142 |
| Western | 0.38 (0.21-0.69) | **0.002** | 0.31 (0.17-0.56) | **<0.001** |
| North Eastern | 0.43 (0.20-0.89) | **0.024** | 1.93 (0.85-4.40) | 0.118 |
| Central | 2.66 (1.48-4.77) | **0.001** | 1.44 (0.97-2.14) | 0.074 |
| **Occupation** |  |  |  |  |
| Unemployed/Unpaid | 1 |  | 1 |  |
| Self-Employed | 1.12 (0.78-1.59) | 0.542 | 0.99 (0.61-1.62) | 0.98 |
| Employed/Salaried | 3.17 (2.02-4.97) | **<0.001** | 1.63 (0.90-2.96) | 0.105 |
| **Obesity** |  |  |  |  |
| Not obese | 1 |  | 1 |  |
| Obese | 3.82 (2.62-5.56) | **<0.001** | 2.01 (1.34-3.01) | **0.001** |
| **Hypertension** |  |  |  |  |
| No hypertension | 1 |  | 1 |  |
| Hypertension | 2.51 (1.86-3.39) | **<0.001** | 1.47 (1.06-2.04) | **0.023** |
| **Model goodness of fit** | **F(9190)=0.72,** | **p=0.693** |  |  |

Notes: cOR = Crude odds ratio; aOR = Adjusted odds ratio

On adjusting for all the covariates, older age (60-69 years: aOR= 4.20, 95% CI: 2.63-6.71; 45-59 years: aOR= 5.57, 95% CI: 2.92-10.65), poorest wealth quintile (aOR= 10.03, 95% CI: 4.77-21.09) and tertiary education (aOR= 3.58, 95% CI: 1.71-7.47) remained the strongest predictors of diabetes screening. The odds of testing increased with increasing age and reducing wealth status. Obese (aOR= 2.01, 95% CI: 1.34-3.01) and hypertensive (aOR= 1.47, 95% CI: 1.06-2.04) participants also had increased odds of diabetes testing. The Western region (aOR= 0.31, 95% CI: 0.17-0.56) had lower odds of diabetes testing compared to the Rift Valley region.

#### Model Diagnostics

The goodness of fit test(p=0.693) showed that the model fits the data adequately and is reliable in estimating the associations between the explanatory variables and diabetes screening. **(Table 2)**

## Discussion

This study found that a small proportion of adults in Kenya have ever been tested for diabetes. Testing was lower among younger adults, individuals with no formal education, the unemployed, those in the highest wealth group, and residents of the Northeastern region. More than half of diabetes cases were undiagnosed, particularly among younger adults, those with primary-level education, individuals in the highest wealth quintile, and people from the Northeastern region. Diabetes prevalence was relatively low, but there was substantial attrition across the care cascade, with notable losses from awareness to treatment and from treatment to glycaemic control. Higher odds of testing were observed among older adults, individuals with higher levels of education, those in lower wealth groups, and those with hypertension or obesity.

The prevalence of diabetes screening in Kenya was quite low (only 11%), while the proportion of undiagnosed diabetes was very high. This finding is consistent with a study of 12 countries in SSA that also revealed a similar trend(39). This finding reflects barriers to accessing diabetes screening and diagnostic services in low resources settings like Kenya. The burden is driven by systemic and resource factors such as limited funding (40)(41,42), low public awareness(43), inadequate workforce capacity (40), shortages of diagnostic tools(44), and limited access to screening facilities(45–47). Efforts to improve diabetes detection in Kenya should prioritise the scale-up of screening services through primary healthcare (PHC) platforms, health system capacity strengthening and raising awareness.

The diabetes care cascade shows substantial attrition and missed opportunities for care across the continuum, consistent with evidence from similar settings in Africa (31,48,49). These losses reflect critical gaps in diagnosis, linkage to care, treatment initiation, and retention(29). Retention in care for diabetes is generally low in Africa(50), with studies reporting low rates of treatment initiation after screening(51) and loss to follow-up after treatment initiation(52). Moreover, treatment coverage has not kept pace with rising diabetes prevalence(53). Gaps in guidelines, coordination, and follow-up result in fragmented diabetes care and limit the health system’s ability to initiate and sustain treatment(42,46). Limited availability of essential medicines, along with high out-of-pocket costs, further hinder treatment access and adherence in LMICs(47,54)(55,56). In Kenya, diabetes patients still incur a very high cost in managing diabetes(57). The diabetes care cascade can be strengthened through improved linkage to care, better access to treatment, and enhanced retention.

In this study, diabetes testing was significantly associated with older age, a finding that has also been reported by other previous studies in Asian populations(41), Peru(58), Lesotho(59) and Nigeria(60). T2DM affects many older adults(61,62), and from our study, the prevalence of diabetes is highest among older people compared to younger individuals. Older adults are more likely to undergo diabetes screening due to greater health awareness and more frequent contact with health services, while younger individuals are less likely to be screened, partly due to the perception that diabetes primarily affects older people (41). Obesity and hypertension were associated with increased odds of testing in the present study. Previous studies have also reported similar observations for obesity(63)(64,65) and hypertension(58,66). Obesity is a key risk factor to diabetes and together with hypertension and other risk factors increases the risk of diabetes complications(67). Individuals under treatment for hypertension are also likely to be targeted for diabetes screening as they are at increased risk(68). The value of screening high risk populations including the obese, hypertension and older adults has been adequately demonstrated(69–71). Diabetes screening in Kenya should prioritize high-risk groups, especially older adults and those with obesity or hypertension, through integrated chronic disease care, while also improving awareness and early detection among younger populations.

Lower wealth status was associated with increased odds of diabetes testing in Kenya contrary to other previous studies that have reported that people with lower wealth status were less likely to participate in diabetes screening(64)(60)(66). The finding in Kenya is also contrary to the global trend where poverty typically impedes healthcare access (72,73). This can be attributed to targeted public health interventions and the underlying disease burden among the low resource communities. Evidence in Kenya further indicates a significant need in poor settings like Kibera Slums where studies have documented a diabetes prevalence of 5.3%(74). In our study, the prevalence of diabetes is highest among the poor, necessitating screening services among this group. In Kenya, proactive, community-based screening approaches led by the Ministry of Health and partners have improved access among underserved populations by reducing barriers such as transport costs, reflecting effective targeting of high-burden groups(24,51,75–78).The finding reflects the success of strategic healthcare delivery in response to a health burden within vulnerable populations.

This study is among the first to examine diabetes testing and care cascade in Kenya. The strength of this study lies in the exploration of diabetes testing, care cascade and simultaneous comparison with diabetes prevalence and undiagnosed diabetes. The analysis is based on a nationally representative survey making the findings generalisable to Kenya. However, this most recent such dataset dates from a survey conducted in 2015. The relatively small size of the dataset is also a limitation, as it constrained the scope of analyses that could be undertaken. In particular, it limited the ability to conduct more detailed statistical analyses, such as regression modelling of outcomes in the later stages of the care cascade. Also, screening has been analysed based on self-report and may be subject to recall bias. Despite these limitations, the study provides important preliminary evidence on diabetes screening and care progression, with findings that are consistent with those reported in similar settings.

## Conclusion

This study highlights low prevalence of diabetes testing alongside a high proportion of undiagnosed diabetes in Kenya, with substantial attrition across the care cascade. Older adults, obese, hypertensive and individuals with low wealth status were more likely to have ever been tested. These results reflect a successful targeting of high burden groups and vulnerable populations with significant gaps in ongoing care. Importantly, there is a need to sustain the observed success while strengthening risk-based screening targeting older adults and people living with obesity and hypertension, while improving linkage and retention in care to improve diabetes outcomes in Kenya.

## Acknowledgement

We would like to thank the Wellcome Trust for funding this study and the Kenya National Bureau of Statistics for granting us access to the data.

## Data availability

Data used for this study are available to download from the Kenya Nationional Data Archive (https://statistics.knbs.or.ke/nada/index.php/catalog/24) upon request from Kenya National Bureau of Statistics.

## Author Contributions

CA conceptualised the study. CA obtained the data and analysed it with input from PJD, CVT, JOO and EOAW. CA drifted the initial manuscript that was reviewed by all the authors and approved for publication.

